# Implementation of a Goals-of-Care Communication Priming Intervention Tailored to Outpatient Stroke Survivors: A Pilot Study

**DOI:** 10.1101/2024.08.04.24311479

**Authors:** Nauzley C. Abedini, Erin K. Kross, Ruth A. Engelberg, Gigi Garzio, Claire J. Creutzfeldt

**Affiliations:** Division of Gerontology and Geriatric Medicine, Department of Medicine, University of Washington, Seattle, Washington, USA; Cambia Palliative Care Center of Excellence, UW Medicine, Seattle, Washington, USA; Division of Pulmonary, Critical Care, and Sleep Medicine, Department of Medicine, University of Washington, Seattle, Washington, USA; Department of Neurology, University of Washington, Seattle, Washington, USA

**Keywords:** goals of care, stroke, caregivers, communication

## Abstract

**Background:** Goals-of-care conversations (GOCC) are important but infrequent after stroke. Serious illness communication priming guides like the Jumpstart Guide can increase GOCC, but have not been evaluated in the stroke population.

**Methods:** We conducted a randomized pilot study to evaluate feasibility and acceptability of the Jumpstart Guide adapted for outpatient stroke survivors, their surrogates, and clinicians. We recruited stroke survivors ≥60 years presenting for care at a single academically-affiliated stroke clinic. We enrolled surrogates if the patient had communication or cognitive impairment. Patients/surrogates were randomized to intervention (patient/surrogate and clinician received pre-visit Jumpstart Guide) or control arms. We assessed feasibility of participant enrollment, survey completion and extraction of GOCC documentation from the electronic medical record. We assessed acceptability using patient/surrogate and clinician surveys.

**Results:** We enrolled 15/24 (63%) eligible patients or surrogates. We randomized 5 patients alone and 3 patients with surrogates to the intervention arm, and 5 patients alone and 2 patients with surrogates to the control arm. Six clinicians were enrolled for the 8 intervention encounters. Patient characteristics in both groups were similar with mean age 74.7 years, 10/15 male, 12/15 white, and 10/15 with acute ischemic stroke. Most patients/surrogates (7/8 intervention vs 7/7 control) and all intervention clinicians completed post-visit surveys. Most intervention participants reported successful pre-visit receipt of the Jumpstart Guide (6/7 patient/surrogates; 6/8 clinicians). Of these, all intervention patients/surrogates and 5/6 clinicians stated they would “definitely” or “probably” recommend it to others. Two intervention vs no control patients had newly documented GOCC post-visit. Intervention patients/surrogates more frequently reported discussing GOCC during their clinic visit (6/7 intervention vs 4/7 control).

**Conclusions:** Implementation of a stroke-specific GOCC priming guide (Jumpstart Guide) in an outpatient stroke clinic is feasible and acceptable. A large randomized controlled trial is needed to evaluate its efficacy in improving outpatient stroke clinic GOCC.

## Introduction

While advances in the prevention and acute management of stroke have substantially reduced the incidence, morbidity, and mortality associated with stroke in the United States (US),^1^ stroke is still the fifth leading cause of death in the US.^2^ Those most severely affected by stroke survive only with the provision of artificial life support, such as mechanical ventilation and artificial nutrition and hydration (ANH). Resultant functional and cognitive impairments can significantly impact quality of life for stroke survivors and their families.^3, 4^ Goals-of-care conversations (GOCC) among clinicians, patients and surrogates aim to clarify patient values and preferences around their medical care so that people receive care that aligns with what is most important to them. GOCC can improve quality of life, improve patient and family assessments of quality of care, and reduce intensity of end-of-life care among seriously ill patients.^5, 6^

Despite a desire from stroke survivors and surrogates to have GOCC^7, 8^ and recommendations from the American Academy of Neurology^9^ and the American Heart Association/American Stroke Association^10^ in support of early GOCC, these conversations are infrequent or inadequate among people affected by stroke.^7, 8^ Most people presenting acutely with stroke have not had prior GOCC, and if they have, they are often not applicable to their current situation.^8, 11, 12^ Among racially and ethnically minoritized stroke survivors,^13–15^ GOCC occur less often.^16^ Therefore, interventions to improve GOCC among stroke survivors and their families must incorporate strategies that enhance inclusion and reduce disparities.

The Jumpstart Guide is a GOCC-priming guide that aims to improve GOCC by (1) prompting the occurrence and documentation of GOCC for patients with chronic or serious illness, and (2) providing clinicians, and in some cases, patients and their surrogates, with specific language to facilitate GOCC. For hospitalized patients and outpatients with chronic, life-limiting diseases, the use of the Jumpstart Guide compared to usual care increased the documentation of GOCC^17, 18^ and, in the outpatient setting, improved patient rating of the quality of communication.^17^ Communication priming guides have not been studied in people affected by stroke, nor have these guides been adapted for use with racially, ethnically, or culturally diverse populations.

Given these gaps, we conducted the second of a 2-phase study aiming to apply human-centered design (HCD) – a strategy that actively engages end-users in the design and implementation of new interventions that we have utilized to adapt the Jumpstart Guide to other populations^19, 20^ – to adapt and pilot the Jumpstart Guide with diverse stroke survivors and clinicians.^21^ HCD’s main characteristics include (1) *insight* - developing an understanding of stakeholders and their needs; (2) *ideation* - engaging stakeholders throughout the design process; and (3) *implementation* of the intervention.^22^ Throughout the HCD process, stakeholder engagement is essential. In the first phase of our study, we conducted qualitative interviews with racially and ethnically diverse stroke survivors, their surrogates, and clinicians, in which we explored their perspectives on facilitators and barriers to outpatient stroke GOCC in general (i.e. *insight*), as well as their perspectives on the proposed stroke-specific Jumpstart Guide (i.e. *ideation*).^21^ These results led to iterative re-design of the stroke-specific Jumpstart Guide tailored to the specific needs of diverse stakeholders. In this manuscript, we describe the second phase of our study in which we conducted a randomized pilot study of our stroke-specific Jumpstart Guide to assess feasibility, acceptability, and preliminary efficacy in improving perceived and documented GOCC in outpatient stroke clinic (i.e. *implementation*).

## Methods

### Participants and Eligibility Criteria

We conducted this pilot trial in the outpatient stroke clinic at a single regional stroke and trauma center affiliated with a large academic health system caring for over 2,000 patients with stroke per year.

#### Patient and Surrogate Eligibility

Two investigators (NCA and CJC) prospectively screened the stroke clinic schedule and electronic health record (EHR) 2-4 weeks prior to the clinic visit to identify stroke survivors presenting for their initial post-stroke consultation with any stroke neurologist between June 22, 2023 and September 1, 2023. Stroke survivors were eligible for participation if they were age 60 years and older, referred from within the UW Medicine system, and had confirmed acute ischemic stroke, hemorrhagic stroke, or transient ischemic attack resulting in prior hospitalization as their primary visit diagnosis. Stroke survivors were ineligible if they had an infarct found incidentally on imaging; had a diagnosis of amaurosis fugax or central artery occlusion; had concurrent cancer, which would impact their prognosis and possibly confound GOCC; preferred a language other than English; or were unreachable by phone during study recruitment calls. If patients were unable to communicate (e.g. due to aphasia, cognitive impairment, or other reasons) the patient’s surrogate was invited to participate in the study.

#### Clinician Eligibility

Clinicians were eligible if they were caring for a stroke survivor who was enrolled in the study, if their primary field of practice was in vascular neurology, and if they longitudinally cared for stroke survivors in the outpatient stroke clinic.

### Recruitment, Randomization, and Study Protocol

#### Patients and Surrogates

Study personnel contacted all eligible stroke survivors or their surrogates by telephone 2 weeks prior to their clinic visit for recruitment. Those who were unreachable after up to 10 attempts were deemed ineligible. Those who were reachable were provided a brief study introduction and were consented verbally. Consenting participants were then randomly assigned to the intervention versus enhanced control in a 1:1 ratio using a computer-generated randomization sequence. During this initial contact, all consenting patients or surrogates received the first of two telephonic reminders from study personnel, including the date of the upcoming clinic visit and a request to have a surrogate or support person accompany the stroke survivor to the visit if possible. The control group was considered “enhanced” usual care because they received the same pre-visit reminders as the intervention group.

All consenting patients or surrogates were then sent an introductory packet consisting of a written copy of the consent form along with a written appointment reminder based on their preferred method (either by electronic or ground mail) approximately 1-1.5 weeks prior to their clinic appointment. Intervention patients or surrogates were also sent a copy of a patient- or surrogate-facing Jumpstart Guide to review with the reminder materials. Following this introductory mailing, both intervention and enhanced control groups received the second appointment reminder call from study personnel approximately 3-5 days prior to their scheduled appointment. This reminder similarly provided the upcoming appointment date and encouragement to include a surrogate or support person at their visit if possible. During this second call, intervention patients or surrogates were also asked if they received the Jumpstart Guide; in the event that they had not received it, the Jumpstart Guide was sent again via electronic mail. Patients or surrogates who ultimately did not appear for their appointment or cancelled their clinic visit were disenrolled from the study.

#### Clinicians

All clinicians whose patients were enrolled in the study were also enrolled in the same corresponding arm (intervention vs enhanced control). One clinician could have more than one patient enrolled in the study and could be enrolled in both arms for separate patients. We capped intervention patients for each clinician to no more than 2 per day to reduce study burden. Clinicians were not asked to provide consent given that the Jumpstart Guide promotes conversations that should be standard of care with stroke survivors.

Three days prior to the clinic visit, clinicians who were seeing stroke survivors in the intervention arm received an email with the clinician-facing Jumpstart Guide as a PDF attachment. Study personnel (GG, NCA) also sent a secure EHR message the day of the clinic visit to remind clinicians to check their email and review the Jumpstart Guide prior to the clinic visit. No pre-visit or post-visit communication was performed with clinicians who were seeing patients in the control arm to limit study burden.

### Data Collection

#### Demographic Data

Prior to the clinic visit, two study investigators (NCA, CJC) extracted stroke survivor demographic information (age, gender, race, ethnicity, place of residence, stroke characteristics), and information about prior advance care planning (including previously documented code status, advance directives or Physician Orders for Life Sustaining Treatments, healthcare power of attorney paperwork) from the EHR. We assessed the patient’s level of disability by collecting the modified Rankin Scale (mRS) from the clinic note. If no mRS was documented, we abstracted the mRS from the neurological exam. Surrogate and clinician demographic data (age, gender, race, ethnicity) were obtained via follow-up survey within 1 week of the clinic visit.

#### Patient and Surrogate Surveys

Study personnel (GG) delivered 1-week post-visit surveys to both intervention and enhanced control patients or surrogates electronically. If individuals did not respond, up to 3 total email attempts were made at 1-week intervals. Email non-responders then received up to 3 follow up telephone calls to complete the survey. A small subset of patients or surrogates specified preferring phone contact over email at the outset of the study. For these individuals, study personnel instead made up to 3 telephone calls to complete post-visit surveys. For patients or surrogates in both study arms, the survey included questions assessing whether they had an opportunity to have GOCC, barriers to having GOCC during the visit (using a predefined list, of which respondents could choose multiple answers or write-in a response), and who prompted the GOCC (clinician vs patient/surrogate vs both). For patients or surrogates in the intervention arm only, questions assessed Jumpstart Guide feasibility (i.e., whether they received the Jumpstart Guide before the clinic visit) and acceptability (i.e., whether they would recommend the Jumpstart Guide to other patients/families/clinicians), and also elicited feedback on how to improve the Jumpstart Guide.

#### Clinician Surveys

Clinicians of patients in the enhanced control arm were not surveyed. For clinicians assigned to the intervention arm, survey questions assessed Jumpstart Guide feasibility (i.e., whether the Jumpstart guide was disruptive or helpful during their clinic day), acceptability (i.e. whether they would recommend the Jumpstart Guide to other colleagues), and perceived efficacy (e.g. “Did the Jumpstart Guide help you start a goals-of-care conversation with the patient?”). Similar to the patient and surrogate surveys, the clinician survey included an open-ended invitation to provide feedback on how to improve the Jumpstart Guide.

#### Documented GOCC

We (NA, CJC) reviewed post-visit EHR documentation for all enrolled patients and surrogates within 4 weeks of the clinic encounter to look for evidence of newly documented GOCC either within encounter notes or recently scanned media items.

#### Data Management

All data were collected and stored in RedCap.

### Data Analysis

Results were tabulated and analyzed in Stata version 17.0. Given this was a pilot study focusing primarily on feasibility and acceptability, a sample size calculation was not performed. Feasibility was assessed based on pre-determined thresholds in the following areas: (1) patient or surrogate enrollment rate equal to or greater than 50% of those contacted; (2) reported receipt of the Jumpstart Guide equal to or greater than 75% by enrolled patients or surrogates and clinicians; (3) post-visit survey completion rates equal to or greater than 75% for enrolled patients, surrogates, and clinicians; and (4) ability to extract our primary outcome of interest, newly documented GOCC after the clinic encounter among intervention vs control. Acceptability was assessed by the proportion of intervention participants reporting they would “definitely” or “probably” recommend the Jumpstart Guide equal to or greater than 75%.

### Standard Protocol Approvals, Registrations, and Patient Consents

This study was performed with ethical approval from the University of Washington Institutional Review Board.

## Results

### Participant Enrollment and Encounter Characteristics

#### Patients and Surrogates

During the study period, 341 patients were seen by eligible stroke neurologists in our outpatient stroke clinic. Of the 62 eligible patients identified via electronic health record screening, 24 patients and/or surrogates were available by phone. Of those contacted, 21 (88%) agreed to participate and 15 (63%; 8 intervention vs 7 control) completed study activities. This enrollment rate exceeded the predetermined feasibility threshold of 50%. Six of the 21 consented and randomized participants (2 intervention, 4 control) either canceled or failed to show to their appointment and were withdrawn from the study. Five of the 15 enrolled patients (33%) had a surrogate who enrolled with them (3 intervention vs 2 control; see **Figure 1** for enrollment flow chart). Nine of the 15 visits were conducted in-person (3/8 intervention vs 6/7 control, 60%) and 6 (5/8 intervention vs. 1/7 control, 40%) were conducted via telehealth. Surrogates were present with the patient in 6 of 15 visits (4/8 intervention vs. 2/7 controls, 40%).

**Figure 1:**
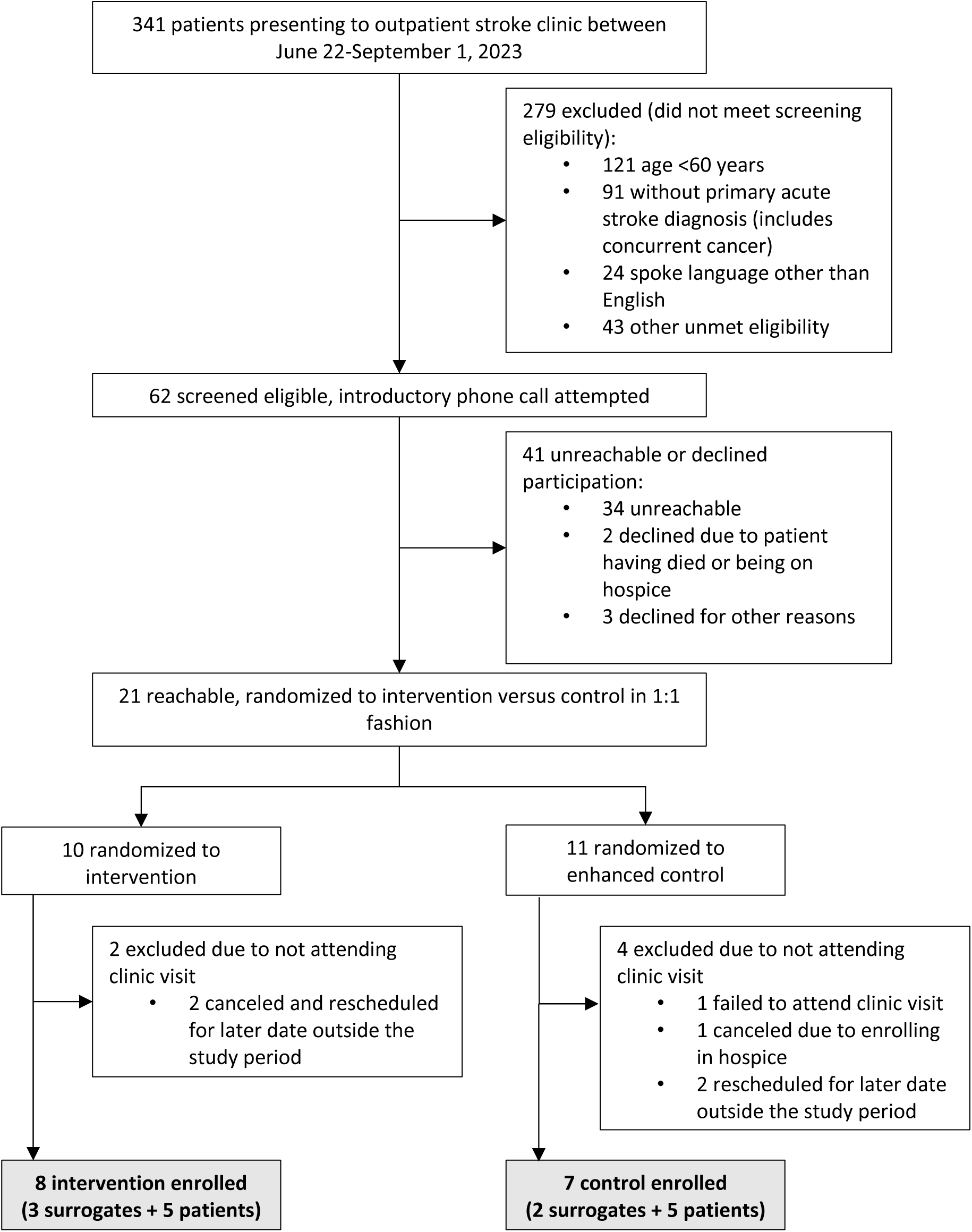
Patient and surrogate enrollment flow chart.

#### Clinicians

Overall, 6 clinicians participated in the intervention arm with 8 intervention patients. Four clinicians had patients both in the control and intervention arms. Of the intervention clinicians, 1 clinician saw 3 intervention patients spread across 2 different clinic days 2 weeks apart.

### Participant Characteristics and Prior Advance Care Planning

We found no major demographic differences between patients in the intervention and control arms. Patients had a mean age of 74.7 years (range 63-83). The majority were male (10/15, 67%), white (12/15, 80%), and had suffered acute ischemic stroke (10/15, 67%) (**Table 1**). Most had strokes involving a single hemisphere (13/15, 87%), and 2 suffered a recurrent stroke (13%). The level of disability was generally mild at time of the clinic visit, with a mean mRS of 1.3 (range 0-3) (**Table 1**). Most of the surrogates identified as female (3/5, 60%) and white (4/5, 80%) and in the age category of 40-59 years (4/5, 80%); one surrogate was over 60 years old.

**Table 1:**
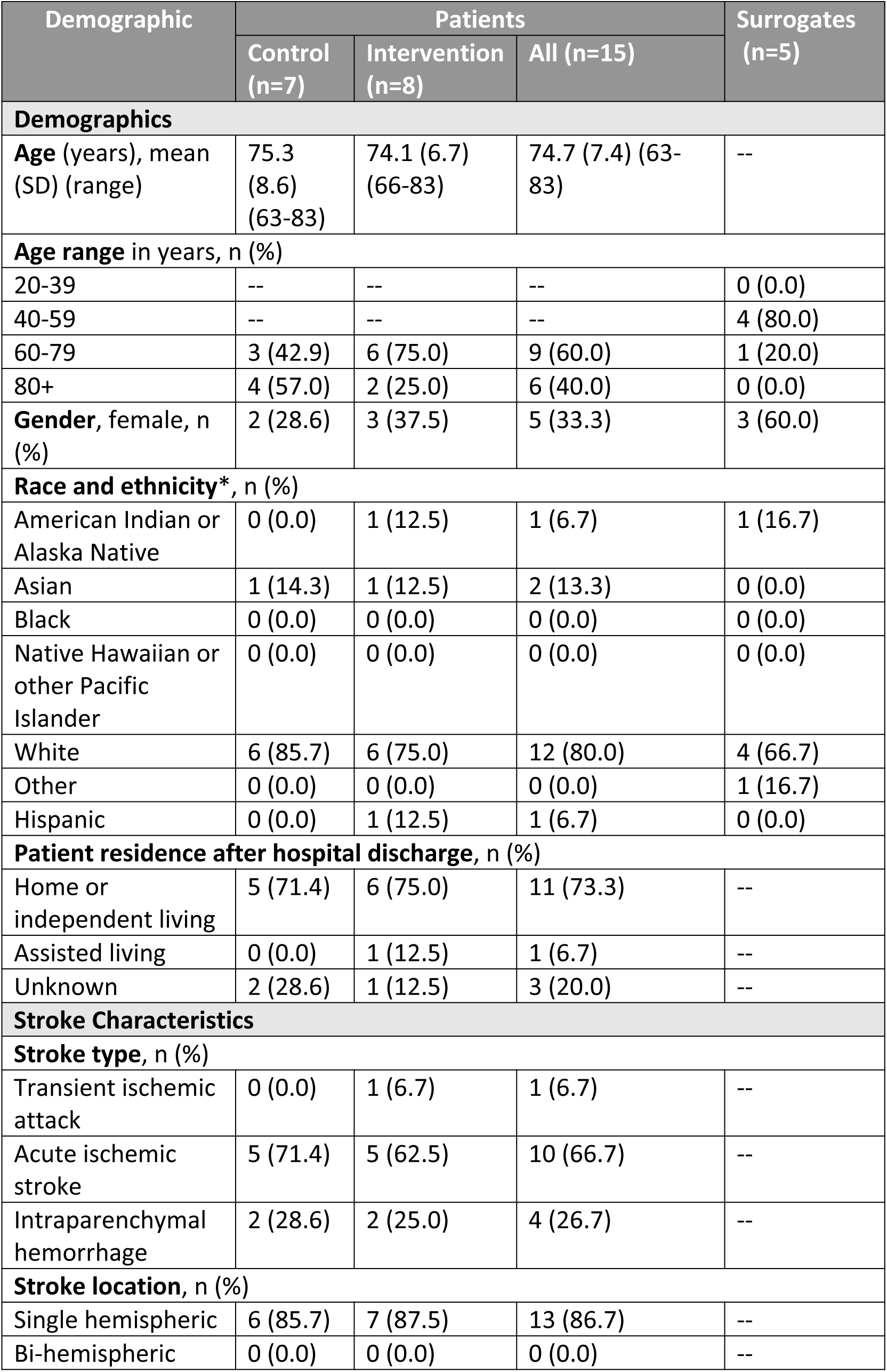
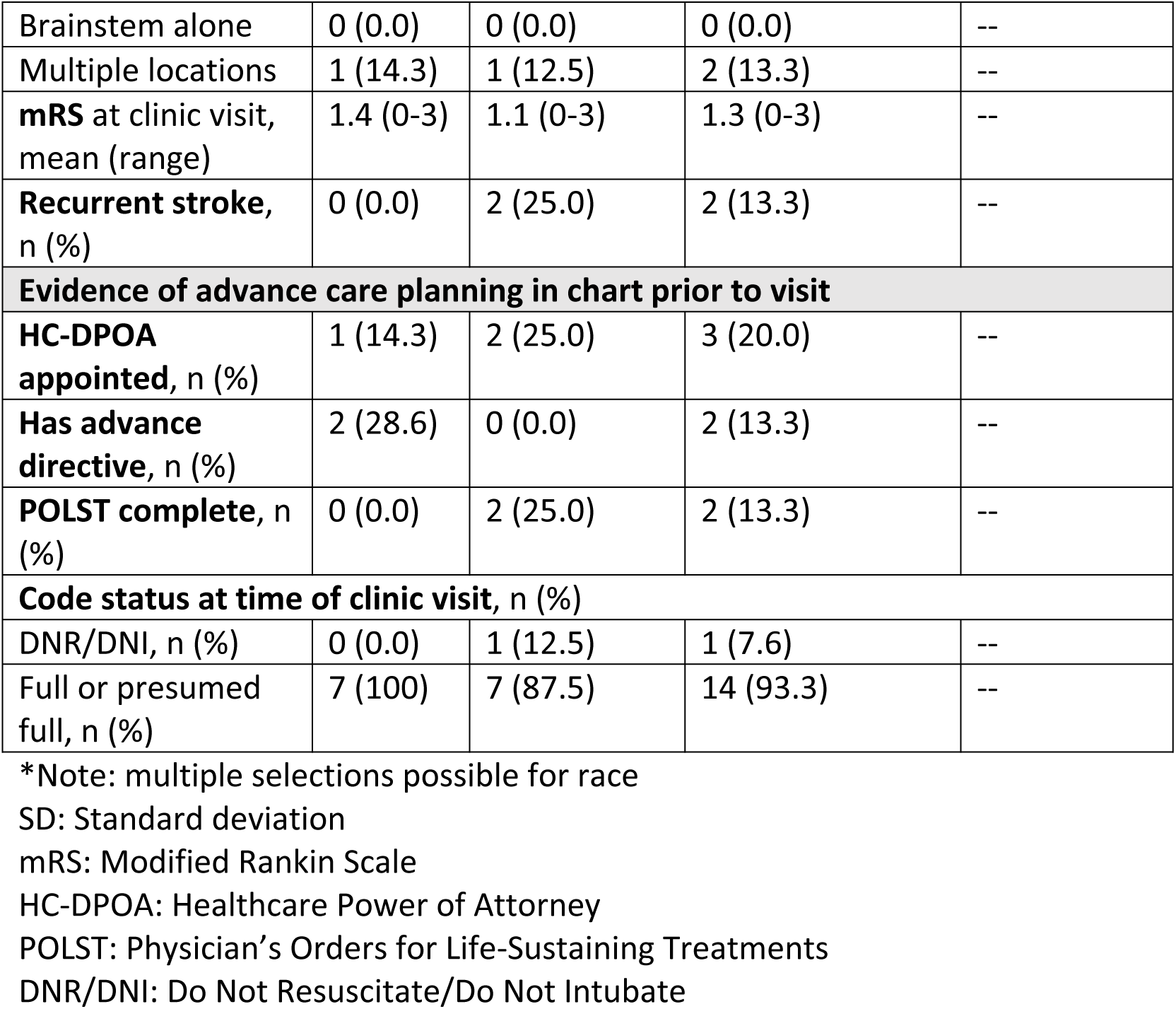
Patient and Surrogate Demographics and Characteristics.

Review of the electronic health record revealed low rates of documented advance care planning prior to the neurology encounter and no major differences between the intervention and control arms. Only 3/15 enrolled patients (20%) had previously appointed a healthcare durable power of attorney (HC-DPOA), 2 (13%) had advance directives, and 2 (13%) had Physician Orders for Life Sustaining Treatments (POLST) uploaded to the chart. Fourteen of 15 patients (93%) had a chart documentation of full or presumed full code (**Table 1**).

Of the clinicians, 4/6 (67%) identified as male 3/6 (50%) were 20-39 years old, and3/6 (50%) were 40-59 years old. Half of the clinicians (3/6) identified as non-White or mixed-race (**Table 2**).

**Table 2:**
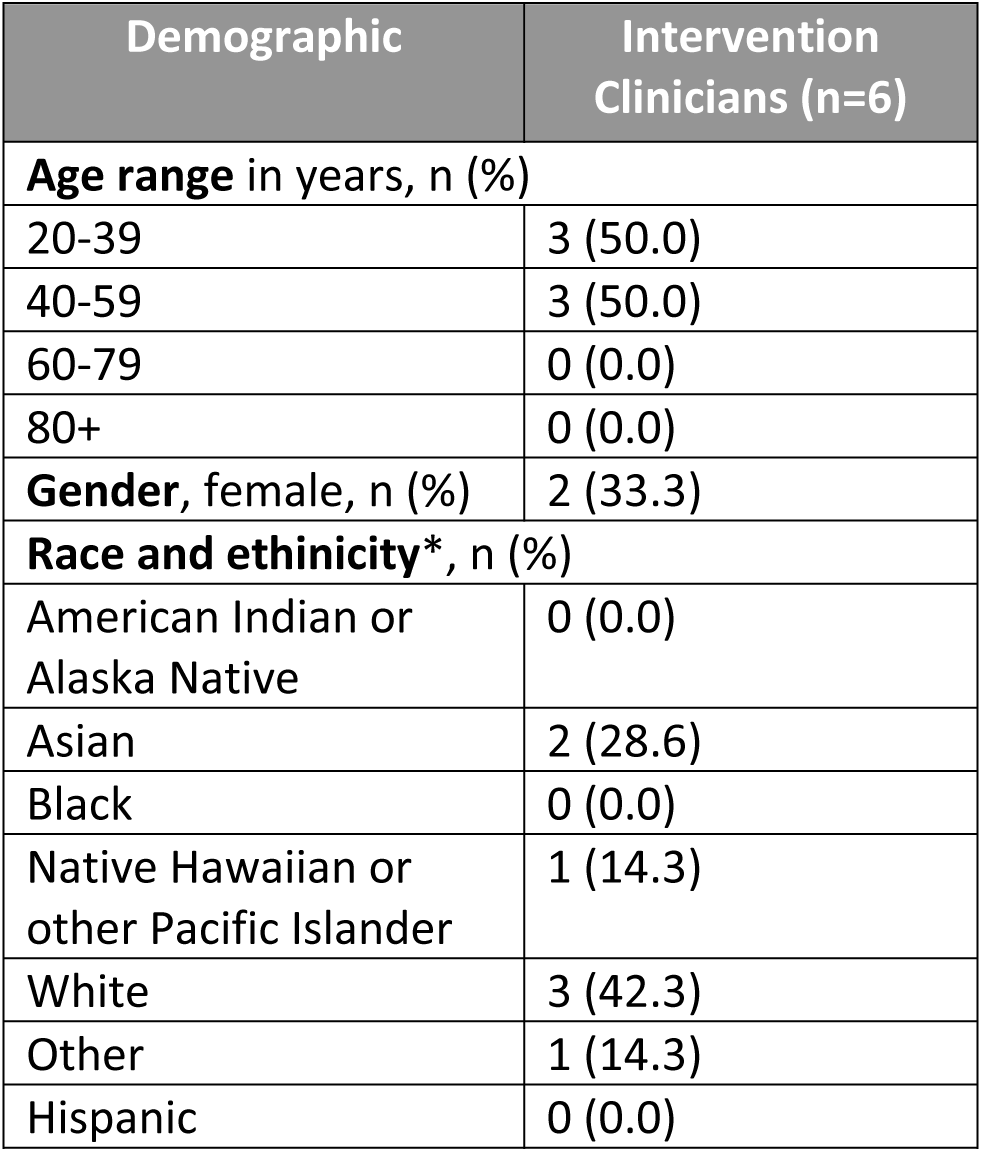
Intervention Clinician Demographics and Characteristics.

### Feasibility and GOCC Outcomes

Fourteen of 15 patients and/or surrogates (93%) completed post-clinic electronic or phone-based surveys (7/8, 88% intervention vs 7/7, 100% control). Clinician post-intervention surveys were completed for all 8 visits by all 6 clinicians (100%) in the intervention arm. In the intervention arm, 6/7 patients or surrogates (86%) and 6/8 clinicians (75%) reported successful receipt of the Jumpstart Guide prior to the clinic visit. Post-survey completion and reported receipt of the Jumpstart Guide met or exceeded the 75% threshold for feasibility set for this study.

Evidence of new GOCC documentation in the electronic health record was found for 2/8 (25%) intervention patients and none of the enhanced control arm patients. A higher proportion of intervention patients and surrogates reported conversations about “goals for future medical care” during their clinic visit (6/7, 86%) as compared with those in the control arm (4/7, 57%). Of those who reported a conversation, more intervention patients and surrogates reported independently initiating the GOCC with their clinician during the visit (3/6, 50%) than did controls (1/4, 25%). GOCC with friends/family after the clinic visit occurred in 4/7 patients in the intervention group and 5/7 in the control group. Four of 6 patients (67%) and surrogates and 2/6 of clinicians (33%) stated that the Jumpstart Guide “probably” helped start a GOCC with the clinician. Two of 6 patients and surrogates (33%) and 4/6 clinicians (67%) felt it “probably” did not help.

### Acceptability

Of those who received the Jumpstart Guide prior to the clinic visit and completed their follow-up survey, 3/6 (50%) patient and surrogates and 1/6 (17%) of clinicians stated they would “definitely” recommend the Jumpstart Guide to others, and 3/6 (50%) of patients and surrogates and 5/6 (83%) clinicians stated they would “probably” recommend it. No clinician felt that the Jumpstart Guide was “disruptive to their clinic day”; 2/6 (33%) found it “very helpful” (corresponding to the 2 cases in which GOCC were also documented in the EHR); 1/6 (33%) found it “somewhat helpful,” and 3/6 (50%) found it “neither disruptive nor helpful” to their clinic day.

### Barriers to GOCC

In reply to an optional question assessing barriers to GOCC in which respondents could choose multiple options, one intervention surrogate noted a lack of time as the main barrier to having a GOCC, whereas 2 patients/surrogates from the control arm offered that they had previously discussed GOCC and were not inclined to readdress them during the visit. Clinicians cited a lack of time as the main barrier in 6/8 instances (75%). One clinician stated that GOCC were previously discussed and did not feel inclined to readdress. Notably, no clinicians felt that discussing GOCC was inappropriate or that they were not comfortable/skilled in this area.

## Discussion

Implementation of a stroke-specific Jumpstart Guide was both feasible and acceptable among stroke survivors, surrogates, and clinicians in an outpatient academically-affiliated stroke clinic. Our study met all pre-defined criteria for feasibility, including: patient or surrogate enrollment rate exceeding 50%; and reported receipt of the Jumpstart Guide and post-visit survey completion rates equal to or greater than 75% among patients, surrogates, and clinicians. While our pilot study was not powered to assess preliminary efficacy, we were able to successfully extract our primary outcome measures of interest, including documented GOCC during the clinic encounter of interest. We also met our primary acceptability criterion in that all intervention participants (patients, surrogates, and clinicians) reported they would “probably” or “definitely” recommend the Jumpstart Guide.

Our finding that few patients in our study had documented GOCC in their charts both prior to the clinic encounter and afterwards is in line with prior findings that GOCC documentation is infrequent for most stroke survivors.^7, 8^ Many reasons may influence this. For patients and surrogates, there may be a lack of perceived urgency,^7^ perhaps related to poor understanding of future stroke risk and overall prognosis.^8^ For clinicians, the high prognostic uncertainty associated with stroke^23^ may contribute to clinician discomfort^7^ around initiating GOCC. Stroke clinicians have also cited perceptions that such discussions may damage the patient-clinician relationship,^7^ and that such conversations may not fall within the purview of their responsibilities when they function as consultants.^21^ Additionally, systematic barriers exist to having GOCC, such as a lack of access to interpreters or inadequate time to conduct such conversations, as has been noted by clinicians in this study and our prior HCD work.^21^ However, most stroke survivors and their family members desire these conversations,^8^ and barriers are frequently clinician-driven. In general, traditional models of GOCC place the responsibility of initiating such conversations in the hands of clinicians. The bi-directional nature of the Jumpstart Guide has the potential to circumvent these barriers so that patients may find the agency to start these important conversations on their own terms. A large, randomized controlled trial is needed to show the efficacy of the Jumpstart guide in improving GOCC in stroke outpatient clinics.

In this pilot study, more patients and surrogates reported that they had a GOCC during their clinic encounter than did clinicians, suggesting that these stakeholder groups may have different notions of what constitutes GOCC, which ultimately may undermine efforts to align values and priorities around future medical care.^24^ Among front-line clinicians, the term “goals of care” can often be ambiguous and inconsistently used, frequently conflated with end-of-life planning or major care transitions, rather than a broader discussion of a patient’s background, values, and perspectives on current and future health states.^25^ We previously found through our HCD that many clinicians equate GOCC with finalizing preferences around life-sustaining treatments and completing documentation (such as a POLST form) to reflect these choices.^21^ In this previous study, many clinicians also likened outpatient GOCC to those held in the inpatient setting, in which conversations may be intensive, long, and usually isolated experiences, rather than a longitudinal, fluid conversation that evolves with the patient as they progress through their illness journey.^21^ This points to another possible benefit of interventions like the Jumpstart Guide in reframing GOCC to be less focused on decisions or concrete outcomes (e.g. changing code status or making decisions around life-sustaining treatments) and instead take a longitudinal and iterative approach to reassessing patient values in the context of their ever-changing health status and uncertain prognosis.

The misperceptions around how much time is required for a GOCC and what constitutes GOCC among stroke neurologists may also explain seemingly paradoxical findings in our study – while most clinicians would recommend the Jumpstart Guide and also felt that having GOCC with their patients was appropriate, a large proportion felt the Jumpstart Guide was not helpful in *initiating* a GOCC. The Jumpstart Guide in isolation may not help allay concerns around the time needed to conduct a full-scale discussion with the intent of changing care goals, but as in most systems-based interventions, will require a multi-pronged approach that includes, but is not limited to, clinician education to advance overall approaches and expectations around GOCC.^26^ Such education may go a long way in helping us move away from a one-and-done conversation, and instead promoting a more measured, longitudinal approach to these discussions to improve the frequency and quality of GOCC for stroke survivors.

In addition to our small sample size, our pilot had several limitations. First, our patient and surrogate populations were largely white and non-Hispanic. Notably, in the HCD process guiding the adaptation of our Jumpstart Guide, we over-sampled patients from racially and ethnically diverse backgrounds.^21^ Thus, we expect that the Jumpstart Guide will be well-suited for patients of many backgrounds and hope that a larger trial with additional language options will introduce opportunities to test our intervention among racially and ethnically diverse patients. Second, our study may have had selection bias in that the stroke survivors enrolled in our study generally had low levels of disability as evidenced by low mRS scores and the fact that most were living at home and/or independently and none in a skilled nursing facility. This is a previously reported limitation at our outpatient stroke clinic in that stroke survivors with the highest degree of disability are least likely to present for follow-up.^8^ While having GOCC with stroke survivors with less disability is still important, it may not have appeared as imminently necessary to participating patients, surrogates, and clinicians. The perspectives of those with severe strokes and multimorbidity and therefore high risk of deterioration – those who ultimately would benefit from GOCC more urgently – were likely underrepresented in this pilot. Third, a number of patients failed to attend their appointments, which resulted in high attrition among those who initially consented to participate in our study. Most of these patients rescheduled within the following 3 months. We anticipate this attrition can be mitigated in a clinical trial in which we have a longer follow up period. Finally, given the small number of providers in our stroke clinic, over half of clinicians were enrolled in both intervention and control arms. We attempted to mitigate the possible implications of survey fatigue by not having control arm clinicians fill out surveys. However, it is possible that having patients in both arms could lead to cross-contamination. One indicator of this would have been if we saw equal numbers of patients in both intervention and control arms with evidence of newly documented GOCC after their visit. We did not see this trend, although our sample size may have precluded us from being able to detect this. In an effort to avoid cross-contamination, we hope to further mitigate the risk of cross-contamination by expanding to multiple sites, thereby increasing the number of eligible clinicians.

In conclusion, our randomized pilot study demonstrates that implementation of the stroke-specific Jumpstart Guide is highly feasible and acceptable among outpatient stroke survivors, surrogates, and clinicians, and may circumvent commonly perceived barriers to GOCC by reframing the content, timeline, and expectations around such discussions.

## Data Availability

Data is stored in RedCap and can be made available upon request.

## Acknowledgements

None

